# Racial and ethnic disparities in COVID-19 disease incidence independent of comorbidities, among people with HIV in the US

**DOI:** 10.1101/2021.12.07.21267296

**Authors:** RA Bender Ignacio, AE Shapiro, RM Nance, BM Whitney, JAC Delaney, L Bamford, D Wooten, M Karris, WC Mathews, HN Kim, SE Van Rompaey, JC Keruly, G Burkholder, S Napravnik, KH Mayer, J Jacobson, MS Saag, RD Moore, JJ Eron, AL Willig, KA Christopoulos, J Martin, PW Hunt, HM Crane, MM Kitahata, E Cachay, for the Centers for AIDS Research Network of Integrated Clinical Systems (CNICS)

## Abstract

**Objectives:** To define the incidence of clinically-detected COVID-19 in people with HIV (PWH) in the US and evaluate how racial and ethnic disparities, comorbidities, and HIV-related factors contribute to risk of COVID-19.

**Design:** Observational study within the CFAR Network of Integrated Clinical Systems cohort in 7 cities during 2020.

**Methods:** We calculated cumulative incidence rates of COVID-19 diagnosis among PWH in routine care by key characteristics including race/ethnicity, current and lowest CD4 count, and geographic area. We evaluated risk factors for COVID-19 among PWH using relative risk regression models adjusted with disease risk scores.

**Results:** Among 16,056 PWH in care, of whom 44.5% were Black, 12.5% were Hispanic, with a median age of 52 years (IQR 40-59), 18% had a current CD4 count < 350, including 7% < 200; 95.5% were on antiretroviral therapy, and 85.6% were virologically suppressed. Overall in 2020, 649 PWH were diagnosed with COVID-19 for a rate of 4.94 cases per 100 person-years. The cumulative incidence of COVID-19 was 2.4-fold and 1.7-fold higher in Hispanic and Black PWH respectively, than non-Hispanic White PWH. In adjusted analyses, factors associated with COVID-19 included female sex, Hispanic or Black identity, lowest historical CD4 count <350 (proxy for CD4 nadir), current low CD4/CD8 ratio, diabetes, and obesity.

**Conclusions:** Our results suggest that the presence of structural racial inequities above and beyond medical comorbidities increased the risk of COVID-19 among PWH

PWH with immune exhaustion as evidenced by lowest historical CD4 or current low CD4:CD8 ratio had greater risk of COVID-19.

## Introduction

The COVID-19 pandemic has had profound direct and structural effects on the health of people living with HIV (PWH) in the US^[1-3]^. Health disparities among Black and Hispanic populations in the US are closely related to structural inequities in social determinants of health^[4]^. The burden of the COVID-19 epidemic in Black and Hispanic communities is the result of deep socioeconomic disparities and inequities that affect health and access to healthcare, increased occupational and household exposure, and physiologic risks resulting from heightened allostatic loads and a disproportionate prevalence of medical comorbidities^[4-10]^.

Black and Hispanic Americans are overrepresented among PWH due to shared structural inequities that also increase risk of HIV acquisition^[11]^. Such inequities may explain, in part, the observations from large population-based studies that showed PWH have a higher risk of severe disease and mortality due to COVID-19 than those without HIV^[12-16]^. Structural inequities in areas of residence, education, and wealth are more prevalent among PWH than in the general population^[17]^, and unevenly distributed by race, ethnicity, and geographic region in the US^[18-20]^.

Little is known about the effect of racial and ethnic disparities on risk of COVID-19 disease in PWH. Structural vulnerabilities among PWH likely contribute to increased risk of exposure to SARS-CoV-2 as well as differential access to testing and healthcare, making infections more or less likely to come to clinical attention. PWH also have a high prevalence of non-HIV-specific comorbidities associated with worse outcomes of SARS-CoV-2 infection. There are significant geographic, racial/ethnic, and income disparities in the distribution of these comorbidities among PWH compared to the general population^[21]^. It is essential to elucidate the role these factors play in COVID-19 incidence, separately from factors that may increase severity of COVID-19 disease in PWH^[22]^. We conducted this study in the longstanding Centers for AIDS Research Network of Integrated Clinical Systems (CNICS) multi-site US cohort of PWH.

## Methods

CNICS is a prospective observational cohort study of adult PWH in routine clinical care at 10 academic institutions across the United States^[23]^. We studied all PWH engaged in care (defined as one or more in-person or virtual HIV primary care visits) between September 1, 2018, and December 31, 2020, and alive on March 1, 2020, at seven participating CNICS sites: Johns Hopkins University, Case Western Reserve University, Fenway Health, University of Alabama at Birmingham, University of California-San Diego, University of North Carolina at Chapel Hill, and University of Washington. CNICS research has been approved by the institutional review boards at each site. Sites were anonymized per regulatory requirements of some sites.

Methods of data collection for the CNICS cohort have been previously reported. Briefly, comprehensive clinical data collected through electronic medical records and other institutional data systems undergo rigorous data quality assessment and are harmonized in a central data repository that is updated quarterly^[23]^. Demographic data, including risk factors for HIV acquisition, are collected at cohort enrollment. Smoking status (current vs. former vs. never tobacco use) was determined through Patient-Reported Outcomes (PRO) collected through tablet-based surveys conducted every 4-6 months in conjunction with primary care visits^[24]^ and supplemented by diagnosis codes.

### SARS-CoV-2 infections

Candidate COVID-19 cases were identified through laboratory test results and provider documented diagnoses (ICD-10) recorded between March 1 and December 31, 2020, and verified through medical record review. SARS-CoV-2 test results and hospitalization records available to CNICS sites from external health systems were reviewed for data completeness. Medical records for all cases identified by diagnosis code without supporting laboratory tests performed within the CNICS system were reviewed by clinicians at each site using a standardized protocol. All appropriately-adjudicated cases were included, regardless of symptom status, which was not always documented for laboratory-diagnosed cases.

### Covariates

We examined demographic covariates (age, sex at birth, self-reported race and Hispanic ethnicity, CNICS site) and the following chronic comorbid conditions: diabetes defined using a previously validated approach as hemoglobin A1c (HbA_1c_) ≥6.5%, a prescription of a diabetes-specific medication, or a diagnosis of diabetes with diabetes-related prescription^[25]^; treated hypertension as a diagnosis of hypertension with prescription of an anti-hypertensive medication; obesity as body mass index (BMI) ≥30 kg/m^2^; hepatitis C virus (HCV) coinfection by the presence of positive HCV antibody or detectable RNA or genotype; and chronic obstructive pulmonary disease (COPD) using a previously validated approach^[26]^ as a diagnosis of COPD and ≥90-day continuous supply of long-acting controller medications. We used laboratory test data to compute clinical measures of chronic kidney disease (CKD) defined as last glomerular filtration rate (eGFR) <60 using CKD-EPI without race adjustment^[27, 28]^, and risk scores for atherosclerotic cardiovascular disease (ASCVD^[29]^) and hepatic fibrosis (Fibrosis-4: FIB-4^[30-32]^). We examined CD4 counts (cells/μL) as both lowest historical value, as a proxy for CD4 nadir, and current CD4 count and CD4/CD8 ratio, as well as HIV viral load (VL; copies/mL). To avoid protopathic bias all laboratory test results were censored one week prior to COVID-19 diagnosis.

### Statistical analysis

We calculated the cumulative incidence and incidence rate of first clinically-detected SARS-CoV-2 infection among PWH by key characteristics from March 1, 2020 through December 31, 2020. SARS-CoV-2 incidence was contrasted by demographic factors including race and ethnicity and geographic location, as well as clinical factors including lowest and current CD4 cell counts. Relative risks for COVID-19 were calculated using relative risk regression^[33]^. We accounted for potential confounding by adjusting models using disease risk scores (DRS)^[34-36]^, the prognostic analogue of propensity scores useful when studying a limited number of exposed patients and outcomes, and a relatively large number of potential confounders (**conceptual model, Supplemental Figure 1**). DRS were constructed independently for each exposure of interest using logistic regression in the full cohort for all non-duplicative covariates^[35]^ (e.g., FIB-4 not adjusted for age, a component of the FIB-4 calculation) including age, birth sex, race/ethnicity, smoking status (current vs. former vs. never use), diabetes, hypertension, and CNICS city. All analyses were conducted in Stata version 17 (StataCorp, College Station, TX).

## Results

Among 16,056 PWH in care during the study period, 44.5% identified as non-Hispanic Black, 37.9% as non-Hispanic White, 12.5% Hispanic, and 5.2% mixed, other, or unreported race/ethnicity. The median age was 52 years (IQR 40-59), 20.9% were female, and 53.6% were MSM. Most (95.5%) were on antiretroviral therapy (ART), and 85.6% had an undetectable HIV VL (**Tables 1, 2**). Overall in 2020, 649 PWH were diagnosed with COVID-19, for a cumulative incidence of clinically-detected infections of 4.0% (95% CI 3.7-4.3). of by the end of 2020. The majority (77%) of COVID-19 cases were confirmed by laboratory testing (SARS-CoV-2 RT-PCR) performed within the CNICS system. The remaining 23% of cases were verified from a variety of sources, including PCR and antigen testing performed in community or public health settings or at non-CNICS-affiliated medical sites. As previously reported, 108 cases (16.3%) were hospitalized, and 12 (1.9%) died^[22]^.

**Table 1.**
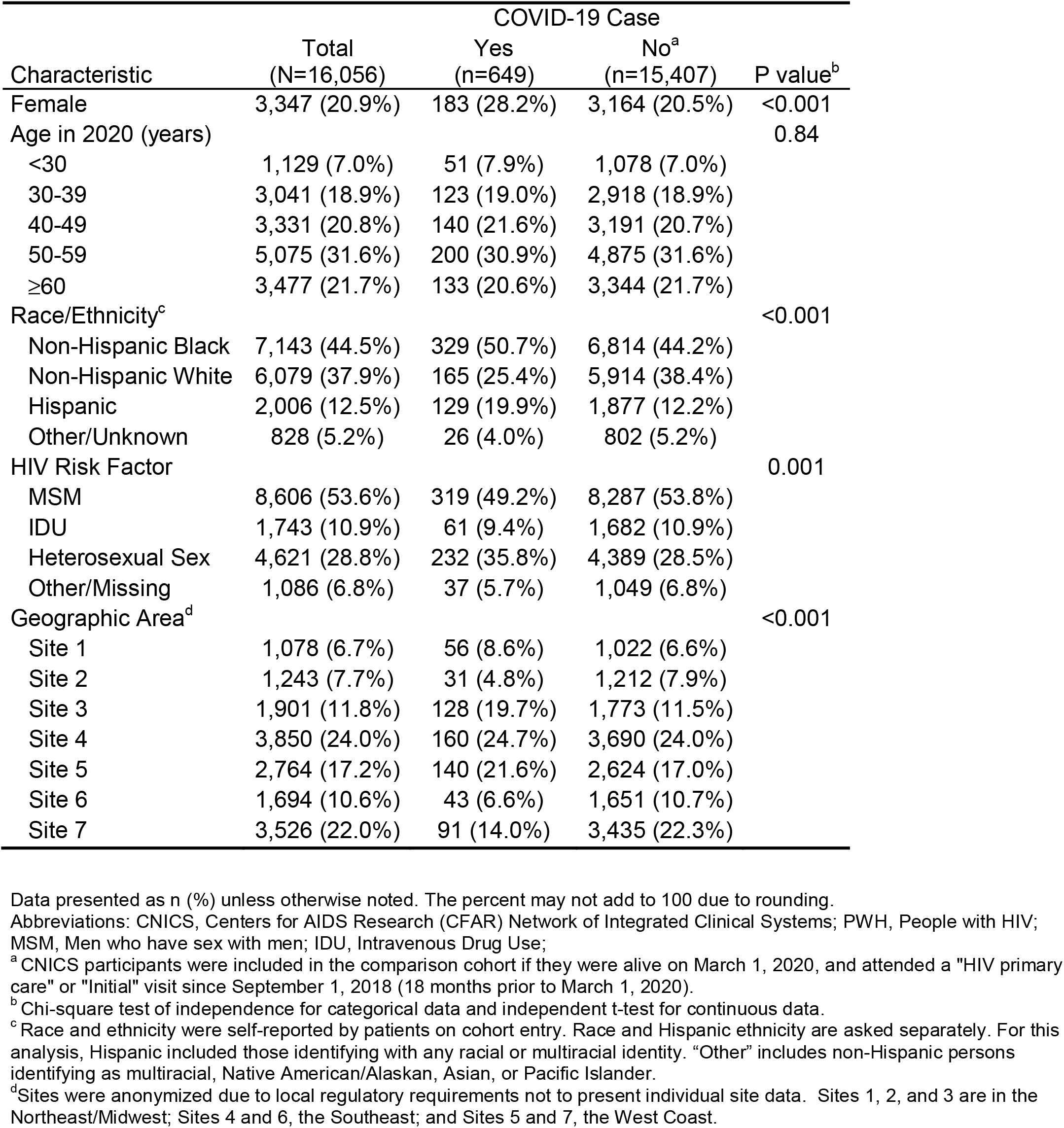
Demographic characteristics of PWH by COVID-19 status.

| Characteristic | Total<br>(N=16,056) | COVID-19 Case |  | P value<sup>b</sup> |
| --- | --- | --- | --- | --- |
|  |  | Yes<br>(n=649) | No<sup>a</sup><br>(n=15,407) |  |
| Female | 3,347 (20.9%) | 183 (28.2%) | 3,164 (20.5%) | <0.001 |
| Age in 2020 (years) |  |  |  | 0.84 |
| <30 | 1,129 (7.0%) | 51 (7.9%) | 1,078 (7.0%) |  |
| 30-39 | 3,041 (18.9%) | 123 (19.0%) | 2,918 (18.9%) |  |
| 40-49 | 3,331 (20.8%) | 140 (21.6%) | 3,191 (20.7%) |  |
| 50-59 | 5,075 (31.6%) | 200 (30.9%) | 4,875 (31.6%) |  |
| ≥60 | 3,477 (21.7%) | 133 (20.6%) | 3,344 (21.7%) |  |
| Race/Ethnicity<sup>c</sup> |  |  |  | <0.001 |
| Non-Hispanic Black | 7,143 (44.5%) | 329 (50.7%) | 6,814 (44.2%) |  |
| Non-Hispanic White | 6,079 (37.9%) | 165 (25.4%) | 5,914 (38.4%) |  |
| Hispanic | 2,006 (12.5%) | 129 (19.9%) | 1,877 (12.2%) |  |
| Other/Unknown | 828 (5.2%) | 26 (4.0%) | 802 (5.2%) |  |
| HIV Risk Factor |  |  |  | 0.001 |
| MSM | 8,606 (53.6%) | 319 (49.2%) | 8,287 (53.8%) |  |
| IDU | 1,743 (10.9%) | 61 (9.4%) | 1,682 (10.9%) |  |
| Heterosexual Sex | 4,621 (28.8%) | 232 (35.8%) | 4,389 (28.5%) |  |
| Other/Missing | 1,086 (6.8%) | 37 (5.7%) | 1,049 (6.8%) |  |
| Geographic Area<sup>d</sup> |  |  |  | <0.001 |
| Site 1 | 1,078 (6.7%) | 56 (8.6%) | 1,022 (6.6%) |  |
| Site 2 | 1,243 (7.7%) | 31 (4.8%) | 1,212 (7.9%) |  |
| Site 3 | 1,901 (11.8%) | 128 (19.7%) | 1,773 (11.5%) |  |
| Site 4 | 3,850 (24.0%) | 160 (24.7%) | 3,690 (24.0%) |  |
| Site 5 | 2,764 (17.2%) | 140 (21.6%) | 2,624 (17.0%) |  |
| Site 6 | 1,694 (10.6%) | 43 (6.6%) | 1,651 (10.7%) |  |
| Site 7 | 3,526 (22.0%) | 91 (14.0%) | 3,435 (22.3%) |  |

Data presented as n (%) unless otherwise noted. The percent may not add to 100 due to rounding.
Abbreviations: CNICS, Centers for AIDS Research (CFAR) Network of Integrated Clinical Systems; PWH, People with HIV; MSM, Men who have sex with men; IDU, Intravenous Drug Use;
<sup>a</sup> CNICS participants were included in the comparison cohort if they were alive on March 1, 2020, and attended a "HIV primary care" or "Initial" visit since September 1, 2018 (18 months prior to March 1, 2020).
<sup>b</sup> Chi-square test of independence for categorical data and independent t-test for continuous data.
<sup>c</sup> Race and ethnicity were self-reported by patients on cohort entry. Race and Hispanic ethnicity are asked separately. For this analysis, Hispanic included those identifying with any racial or multiracial identity. "Other" includes non-Hispanic persons identifying as multiracial, Native American/Alaskan, Asian, or Pacific Islander.
<sup>d</sup> Sites were anonymized due to local regulatory requirements not to present individual site data. Sites 1, 2, and 3 are in the Northeast/Midwest; Sites 4 and 6, the Southeast; and Sites 5 and 7, the West Coast.

**Table 2.**
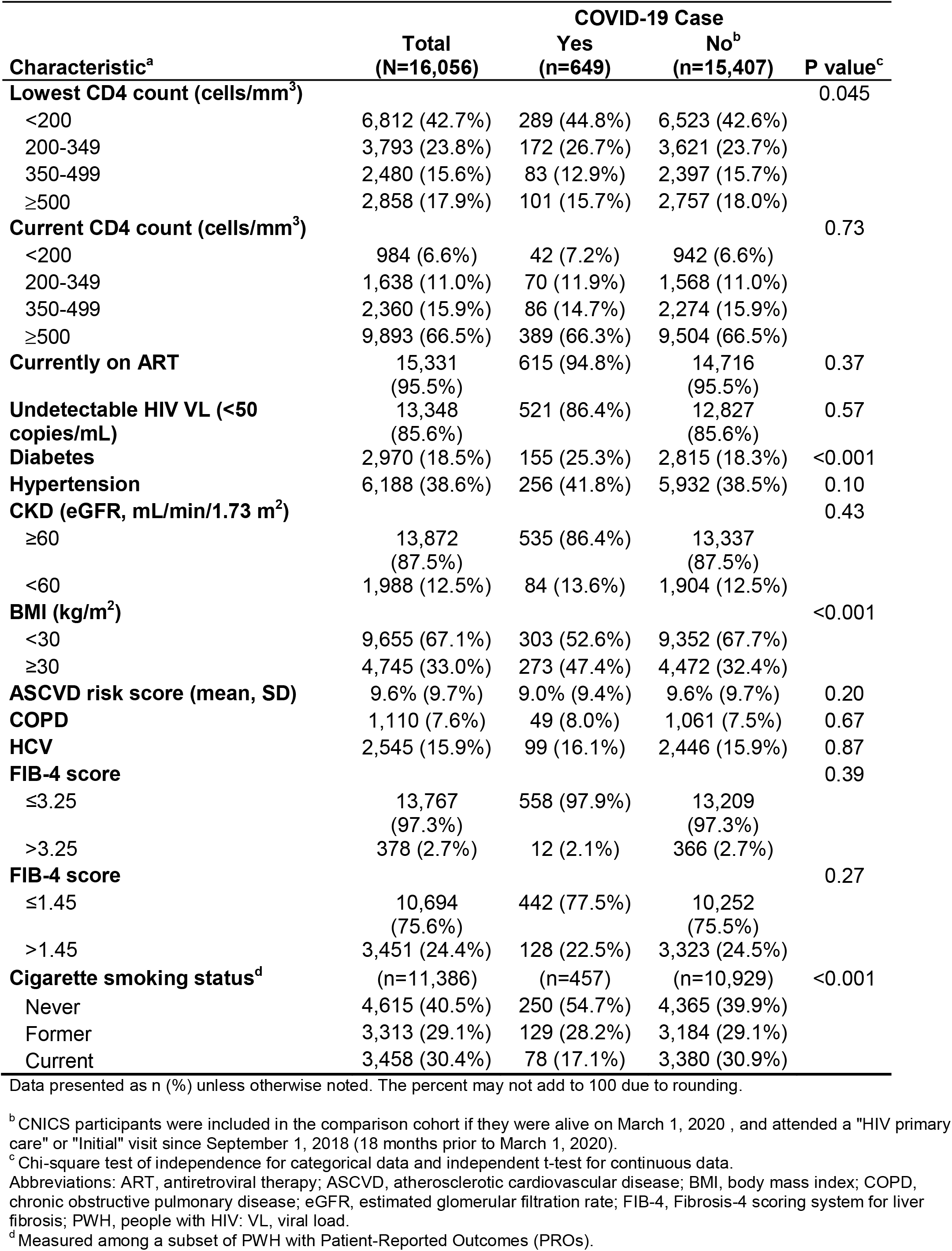
Clinical characteristics of PWH by COVID-19 status.

COVID-19 incidence among PWH increased from 3.29/100 person-years (174 events, 5283 PY) during the second quarter of 2020 to 4.94/100 PYs (95% CI 4.57-5.33) by the end of 2020 By the end of 2020, the cumulative incidence in Black PWH was 5.64/100PY, 7.95/100PY in Hispanic PWH, and 3.30/100 PY for non-Hispanic White (p<0.0001 for both comparisons to non-Hispanic White (**Table 3 and Figure 1**)). Similarly, the IRR for COVID-19 was 1.71 (95% CI 1.42-2.07) for Black and 2.40 (95% CI 1.91-3.01) for Hispanic relative to non-Hispanic White PWH. There were geographic differences in the cumulative incidence of COVID-19 across sites, with cumulative incidence rates ranging from 3.03 to 8.32 per PY by site. COVID-19 cumulative incidence was highest among those with prior low CD4 cell count <350. We did not observe differences according to current CD4 cell count.

**Table 3.** Cumulative Incidence of COVID-19 among PWH (2020).

|  | COVID-19 cases | Person-years<sup>a</sup> | Rate per 100 person-years | P value<sup>b</sup> | P value<sup>c</sup> |
| --- | --- | --- | --- | --- | --- |
| Overall | 649 | 13,130 | 4.94 |  |  |
| Race/Ethnicity<sup>d</sup> |  |  |  |  | <0.001 |
| Non-Hispanic Black | 329 | 5830 | 5.64 | <0.001 |  |
| Non-Hispanic White* | 165 | 5002 | 3.30 |  |  |
| Hispanic | 129 | 1623 | 7.95 | <0.001 |  |
| Other/Missing | 26 | 675 | 3.85 | 0.5 |  |
| Site<sup>e</sup> |  |  |  |  | <0.001 |
| Site 1 | 56 | 880 | 6.37 | <0.001 |  |
| Site 2* | 31 | 1023 | 3.03 |  |  |
| Site 3 | 128 | 1538 | 8.32 | <0.001 |  |
| Site 4 | 160 | 3145 | 5.09 | 0.007 |  |
| Site 5 | 140 | 2249 | 6.23 | <0.001 |  |
| Site 6 | 43 | 1397 | 3.08 | 1.0 |  |
| Site 7 | 91 | 2899 | 3.14 | 0.9 |  |
| Current CD4 count |  |  |  |  |  |
| CD4 <350 | 121 | 2317 | 5.22 | 0.5 |  |
| CD4 ≥350* | 528 | 10813 | 4.88 |  |  |
| Lowest CD4 count |  |  |  |  |  |
| CD4 <350 | 464 | 8728 | 5.32 | 0.006 |  |
| CD4 ≥350* | 185 | 4403 | 4.20 |  |  |

\*Reference group
<sup>a</sup> Person-year time from 03/01/2020 through 12/31/2020.<sup>b</sup> Log-rank test for equality of survivor functions, compared to the reference group.<sup>c</sup> Log-rank test for equality of survivor functions, comparing across all categories.<sup>d</sup> Race and ethnicity were self-reported by patients on cohort entry. Race and Hispanic ethnicity are asked separately. For this analysis, Hispanic includes those identifying as Hispanic with any racial or multiracial identity. "Other" includes non-Hispanic persons identifying as multiracial, Native American/Alaskan, Asian, or Pacific Islander.<sup>e</sup> Sites were anonymized due to local regulatory requirements to not present individual site data. Sites 1, 2, and 3 are in the Northeast/Midwest; Sites 4 and 6, the Southeast; and Sites 5 and 7, the West Coast.

**Figure 1.**
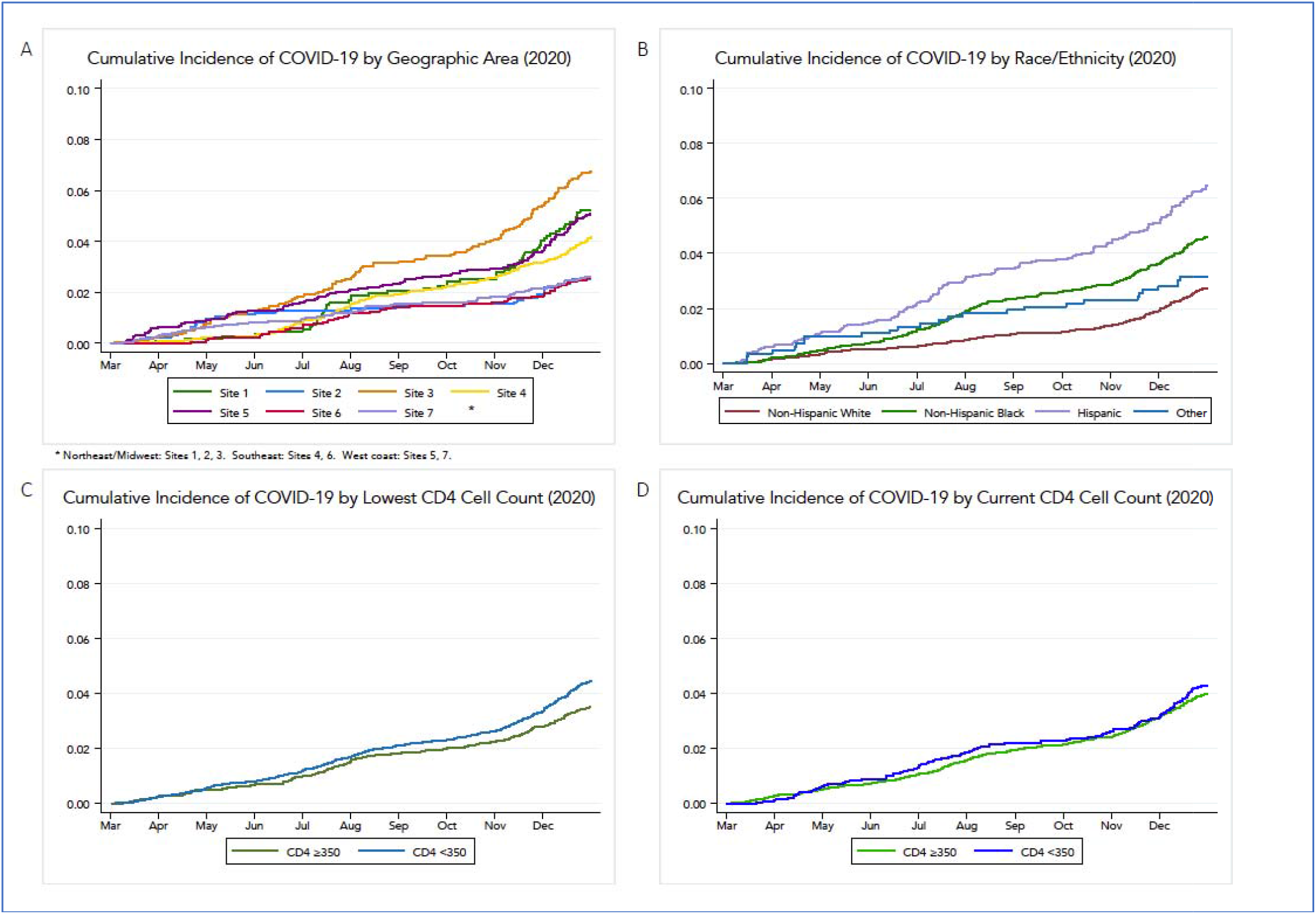
Cumulative Incidence of COVID-19 cases in 2020 by: (A) Geographic Area (B) Race/Ethnicity, (C) lowest historical CD4 T cell count, and (D) current CD4 T cell count. Sites were anonymized based on regulatory requirements of this cohort to not present individual site data. Y axes are cumulative incidences, presented as the proportion of the population under study. Results for current or lowest historical CD4 T cell count wre not different when stratified using 200 cells/mL as the cutoff.

In unadjusted analyses, we observed sociodemographic disparities in risk of COVID-19 also seen in the US general population, namely women and Black and Hispanic PWH were at increased risk of COVID-19 compared to men and non-Hispanic White PWH, respectively (p<0.001 for all). However, older PWH were not at higher risk of COVID-19 (**Table 1)**. PWH who had diabetes or a BMI ≥30 had greater risk of COVID-19 than PWH without those conditions (p<0.001, **Table 2**). We observed a trend towards higher risk of COVID-19 in those with hypertension (p=0.10) and the “smoker’s paradox”^[37]^, with ever-smokers having lower risk of COVID-19 than never smokers (50.2% of cases vs. 55.2% non-cases, p=0.013). CKD, ASCVD risk score, COPD, HCV, and elevated FIB-4 score were not associated with COVID-19 in unadjusted analyses. PWH with a lowest historical CD4 count <350 cell/mm^3^ had a higher risk of COVID-19 than those with a lowest CD4 ≥350 (26.7% and 44.8% of COVID-19 cases vs. 23.7% and 42.6% of non-COVID cases, respectively). The most recent CD4 count measured a median interval of 8.3 (IQR 3.2-19.0) months before COVID-19 diagnosis in cases, was not associated with COVID-19 (p=0.73), nor was ART status or detectable VL, although few in the cohort were not on ART (4.5%) or unsuppressed (15.4%).

In adjusted analyses, racial and ethnic identity was a strong predictor of COVID-19 in PWH (**Figure 2)**. Hispanic (RR 2.03, 95% CI 1.62-2.55, p<0.001) and Black PWH (RR 1.33, 95% CI 1.10-1.61, p=0.003) had the highest risks of COVID-19. Other characteristics associated with COVID-19 after adjustment included female sex (RR 1.32, 95% CI 1.12-1.56, p=0.001), diabetes (RR 1.37, 95% CI 1.15-1.63, p<0.001), and having a BMI ≥30 (RR 1.58, 95% CI 1.36-1.83, p<0.001). The smoker’s paradox remained in adjusted analysis (RR 0.77, 95% CI 0.66-0.89, p=0.001). Neither CKD, HTN, HCV, COPD, nor ASCVD or FIB-4 risk scores predicted COVID-19 diagnosis. PWH whose lowest CD4 count was <350 had significantly higher risk of COVID-19 compared with those with lowest count ≥350 (RR 1.20, 95% CI 1.02-1.42), and we did not find an association with current CD4 count, at either a cut-point of <200 or <350. In addition, PWH with higher current CD4:CD8 ratio had decreased risk of COVID-19 (RR 0.92 for each 1 SD increase in ratio, 95% CI 0.86-0.99, p=0.042). Neither ART status nor detectable VL were predictive of COVID-19 in adjusted analysis (p=0.84 and 0.70, respectively).

**Figure 2:**
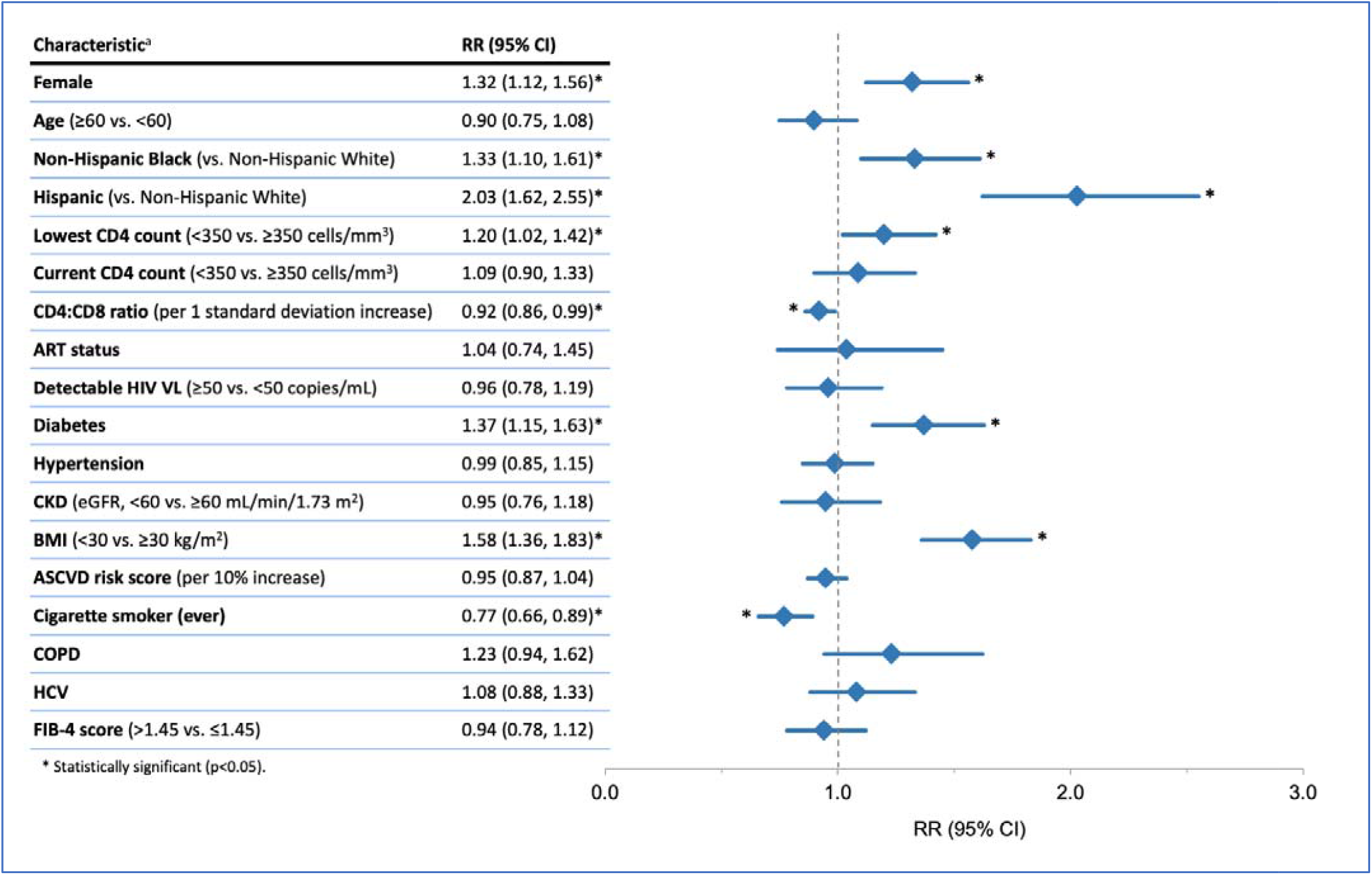
Relative Risk of COVID-19 among PWH by key characteristics. Abbreviations: ASCVD, atherosclerotic cardiovascular disease; BMI, body mass index; CKD, chronic kidney disease; COPD, chronic obstructive pulmonary disease; eGFR, estimated glomerular filtration rate; FIB-4, Fibrosis-4 scoring system for liv r fibrosis; HCV, Hepatitis C virus; PWH, people with HIV; RR, relative risk VL, viral load. ^a^ Relative risk regression models adjusted for demographic and clinical characteristics using disease risk scores, except for the ASCVD risk score analysis which is unadjusted. Disease risk scores were constructed independently for each exposure variable of interest using all non-duplicative covariates.

## Discussion

In this large multi-site cohort of PWH receiving care in the US in 2020 prior to availability of SARS-CoV-2 vaccines, persons who were Hispanic, Black or female had significantly higher risk of COVID-19, as were those who had a lowest historical CD4 count <350 or lower current CD4/CD8 ratio. As seen in the general population and prior analyses of PWH, risk of COVID-19 diagnosis was higher in PWH with obesity and diabetes^[12, 38-40]^. The higher rates of COVID-19 among Black and Hispanic PWH persisted throughout 2020 after adjustment for other clinical and demographic risks. Results of our study in this diverse population of PWH suggest that structural disparities affect COVID-19 incidence above and beyond the disproportionate burden of medical comorbidities. Our findings in PWH, carefully adjusted for measurable demographic and clinical risk factors, expand on observations in the general population that have highlighted how structural inequities in housing, wealth, healthcare access, and occupations are associated with both increased risk of exposure to SARS-CoV-2 and of COVID-19 disease. The nearly three-fold risk difference of being diagnosed with COVID-19 between sites reflects differences in exposure risk and access to testing and care.

The finding that lowest CD4 count <350 cells/mL or low current CD4/CD8 ratio was more predictive of COVID-19 than current absolute CD4 count suggests that HIV-associated immune exhaustion is a driver of symptomatic SARS-CoV-2 infection^[41]^. These results align with other observations showing that SARS-Cov-2 specific T-cell responses correlate with CD4:CD8 ratio and percentage of naïve CD4-T Cells in PWH^[42]^. Neither ART status nor virologic suppression were associated with COVID-19, but few people in our cohort were not on ART or virologically unsuppressed. Notably, we found no association between age and COVID-19, which may reflect stricter adherence to COVID-19 precautions in older adults with HIV. Older PWH who have lived through the first decades of the AIDS pandemic, especially those with personal experience of AIDS, may have been more likely to limit social interactions or request work accommodations during 2020^[43, 44]^. However, once infected, older PWH were more likely to experience severe outcomes of COVID-19 infection^[15, 22]^ as we previously reported ^[22]^.

Limitations of our study include lack of systematic surveillance testing, wherein case identification is limited by access to testing and availability of results, and ascertainment limited to PWH engaged in care. PWH with asymptomatic or minimally symptomatic COVID-19 were likely undertested and under-reported in medical records, a limitation also present in both cohort studies and public health data. Strengths of our study include comprehensive data on more than 16,000 PWH in care across the US with well-characterized comorbidities and HIV-specific clinical data. The latter allowed greater measurement precision to distinguish between the contribution of comorbidities, race/ethnic identity, and HIV factors to risk of COVID-19 diagnosis than studies using public health data. During the earliest months of the pandemic, people with mild symptoms or lacking risk factors for severe disease were discouraged from accessing clinical care, which may have introduced some bias in who was tested during those months^[45]^. Facility-based testing was often performed in emergency departments and hospitals, thus, people with advanced age or comorbidities may have been overrepresented amongst reported cases, although we do not expect that there was differential testing by CD4 count, ART status, sex, or race/ethnicity among people who presented to care. Conversely, persons with a history of AIDS or other comorbidities, who perceived themselves at greater risk of poor outcomes, may have been more likely to test after exposure or with mild symptoms, introducing potential confounding by indication.^[45, 46]^ If present, these biases likely underestimate the effect of racial/ethnic identity on COVID-19 risk, as marginalized populations had less access to testing early in the pandemic, as well as differential health-seeking behaviors due to medical mistrust. While PWH who reported a history of IDU as a risk factor for HIV were not at increased risk of COVID-19, this historical risk category is a poor measure of current high-risk behaviors. We plan future analyses with a subsequent year of COVID-19 cases and PRO data in our cohort to study the effect of substance use including current IDU, and geocoding-based social determinants of health, on COVID-19 risk.

Race and ethnic identity are imperfect, albeit accessible proxies for the experience of structural racism, not directly assessable in this study. Other forms of stigmatization, including anti-LGBTQ bias, also increase allostatic loading^[4, 47-50]^ and compound the structural barriers and health inequities faced by many PWH with marginalized identities. The increased incidence of COVID-19 among PWH with racialized identities and living in certain geographic regions further highlights how disparities in the US collide in the HIV and COVID-19 pandemics^[3]^.

## Conclusions

The results of our study in this diverse US population of PWH suggest that structural disparities differentially impact COVID-19 incidence, with higher and disproportionate rates of COVID-19 in women and Black and Hispanic PWH, and vast risk differences by geography. PWH with lowest CD4 count <350 cells/mL or lower current CD4/CD8 ratio had greater risk of COVID-19. Incidence estimates reflect clinically-detected COVID-19 diagnoses and provide conservative estimates for SARS-CoV-2 infection in the absence of surveillance testing.

## Data Availability

All data produced in the present study are available upon reasonable request to the authors

## Funding and Acknowledgments

We gratefully acknowledge the CNICS cohort participants and their providers for contributing to the study. Funding for this study and for CNICS came from the National Institute of Allergy and Infectious Diseases (NIAID) [CNICS R24 AI067039; UW CFAR NIAID Grant P30 AI027757; UAB CFAR grant P30 AI027767; UNC CFAR grant P30 AI50410; UCSD CFAR grant P30 AI036214 and by the Office of the President of the University of California COVID-19 seed grant; Case Western Reserve University CFAR grant P30 AI036219; Fenway Health/Harvard CFAR grant P30 AI060354, UCSF CFAR grant P30AI027763 and JHU CFAR grant P30 AI094189] and the National Institute on Drug Abuse (NIDA) [R01DA047045].

---

**Supplemental Figure 1:**
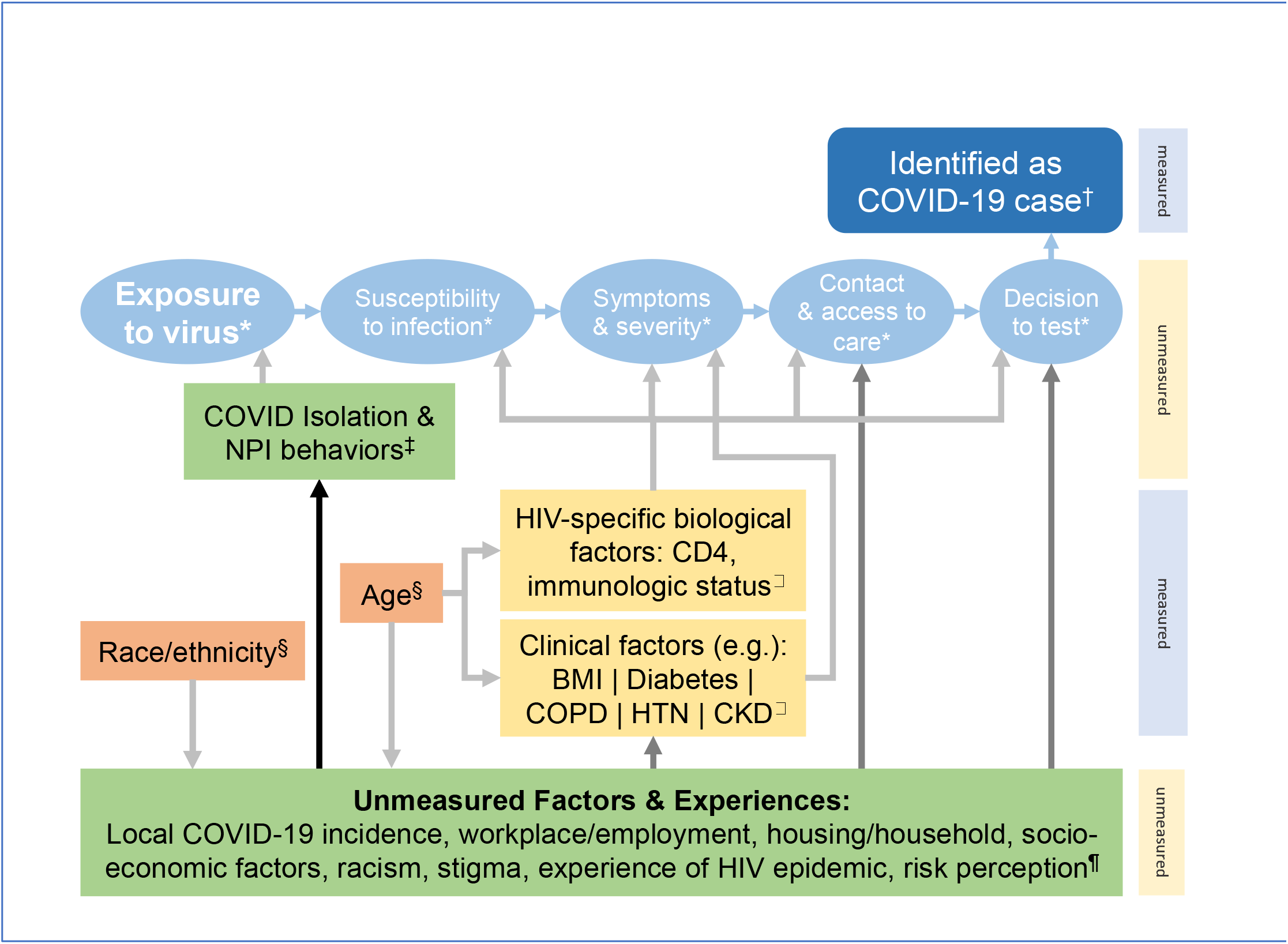
Conceptual Diagram of Factors contributing to identification as a COVID-19 case. Conceptual model of pathway to identification of a case of COVID-19 – simplified * pathway to case identification (unmeasured components of what is ultimately measured) † end of pathway: Case of COVID-19 (measured) ‡ modifiable COVID-related behaviors (unmeasured) § non-modifiable demographic characteristics □ clinical/biological factors (measured) ¶ unmeasured/latent individual properties/experiences that substantially affect a person’s health and behaviors Shade of grey arrows is proportional to degree of effect We employed disease risk scores (DRS) rather than standard multivariate adjustment to prevent overcorrection for colinear effects, especially when attempting to correct for the disproportionate burden of medical comorbidities on Black and Hispanic PWH to evaluate the residual effects of racism/marginalized identities on the association of racial/ethnic identity on COVID-19 diagnosis. Abbreviations: NPI, non-pharmacologic interventions; BMI, body mass index; COPD, chronic obstructive pulmonary disorder; HTN, hypertension; CKD, chronic kidney disease

## References

1. Armstrong WS, Agwu AL, Barrette E-P, Ignacio RB, Chang JJ, Colasanti JA, et al. Innovations in HIV care delivery during the COVID-19 pandemic: Policies to strengthen the Ending the Epidemic Initiative–A Policy Paper of the Infectious Diseases Society of America and the HIV Medicine Association. Clinical Infectious Diseases 2020.

2. Millett GA, Honermann B, Jones A, Lankiewicz E, Sherwood J, Blumenthal S, et al. White counties stand apart: The primacy of residential segregation in COVID-19 and HIV diagnoses. AIDS patient care and STDs 2020; 34(10):417–424.

3. Millett GA. New pathogen, same disparities: why COVID-19 and HIV remain prevalent in US communities of colour and implications for ending the HIV epidemic. Journal of the International AIDS Society 2020; 23(11):e25639.

4. Millett GA, Jones AT, Benkeser D, Baral S, Mercer L, Beyrer C, et al. Assessing differential impacts of COVID-19 on black communities. Ann Epidemiol 2020; 47:37–44.

5. Kullar R, Marcelin JR, Swartz TH, Piggott DA, Macias Gil R, Mathew TA, et al. Racial disparity of coronavirus disease 2019 in African American communities. The Journal of infectious diseases 2020; 222(6):890–893.

6. Khazanchi R, Evans CT, Marcelin JR. Racism, not race, drives inequity across the COVID-19 continuum. JAMA network open 2020; 3(9):e2019933–e2019933.

7. Macias Gil R, Marcelin JR, Zuniga-Blanco B, Marquez C, Mathew T, Piggott DA. COVID-19 Pandemic: Disparate Health Impact on the Hispanic/Latinx Population in the United States. The Journal of Infectious Diseases 2020; 222(10):1592–1595.

8. Bui DP, McCaffrey K, Friedrichs M, LaCross N, Lewis NM, Sage K, et al. Racial and ethnic disparities among COVID-19 cases in workplace outbreaks by industry sector—Utah, March 6–June 5, 2020. Morbidity and Mortality Weekly Report 2020; 69(33):1133.

9. Cordes J, Castro MC. Spatial analysis of COVID-19 clusters and contextual factors in New York City. Spatial and Spatio-temporal Epidemiology 2020; 34:100355.

10. Rodriguez-Diaz CE, Guilamo-Ramos V, Mena L, Hall E, Honermann B, Crowley JS, et al. Risk for COVID-19 infection and death among Latinos in the United States: examining heterogeneity in transmission dynamics. Annals of epidemiology 2020; 52:46-53. e42.

11. Services UDoHH. US HIV Statistics. In; 2021.

12. Yang X, Sun J, Patel RC, Zhang J, Guo S, Zheng Q, et al. Associations between HIV infection and clinical spectrum of COVID-19: a population level analysis based on US National COVID Cohort Collaborative (N3C) data. The Lancet HIV 2021.

13. Braunstein SL, Lazar R, Wahnich A, Daskalakis DC, Blackstock OJ. Coronavirus Disease 2019 (COVID-19) Infection Among People With Human Immunodeficiency Virus in New York City: A Population-Level Analysis of Linked Surveillance Data. Clinical Infectious Diseases 2020; 72(12):e1021–e1029.

14. Durstenfeld MS, Sun K, Ma Y, Rodriguez F, Secemsky EA, Parikh RV, et al. Impact of HIV Infection on COVID-19 Outcomes Among Hospitalized Adults in the US. medRxiv 2021.

15. Bertagnolio S, Thwin S, Silva R, Ford N, Baggaley R, Vitoria M, et al. Clinical characteristics and proctnostic factors in people living with HIV hospitalized with COVID-1 9: findings from the WHO Global Clinical Platform. In: 11th International AIDS Society Conference on HIV Science; 2021. pp. 18–21.

16. Bhaskaran K, Rentsch CT, MacKenna B, Schultze A, Mehrkar A, Bates CJ, et al. HIV infection and COVID-19 death: a population-based cohort analysis of UK primary care data and linked national death registrations within the OpenSAFELY platform. The Lancet HIV 2021; 8(1):e24–e32.

17. Pellowski JA, Kalichman SC, Matthews KA, Adler N. A pandemic of the poor: social disadvantage and the US HIV epidemic. American psychologist 2013; 68(4):197.

18. (CDC). CfDCaP. HIV Surveillance Report. Vol. 31 2018.

19. Dasgupta S, Oster AM, Li J, Hall HI. Disparities in Consistent Retention in HIV Care--11 States and the District of Columbia, 2011-2013. MMWR Morb Mortal Wkly Rep 2016; 65(4):77–82.

20. Reif S, Safley D, McAllaster C, Wilson E, Whetten K. State of HIV in the US Deep South. Journal of community health 2017; 42(5):844–853.

21. Weiser JK, Tie Y, Beer L, Fanfair RN, Shouse RL. Racial/Ethnic and Income Disparities in the Prevalence of Comorbidities that Are Associated With Risk for Severe COVID-19 Among Adults Receiving HIV Care, United States, 2014–2019. Journal of Acquired Immune Deficiency Syndromes (1999) 2021; 86(3):297.

22. Shapiro AE, Bender Ignacio RA, Whitney BM, Delaney JC, Nance RM, Bamford L, et al. Factors associated with severity of COVID-19 disease in a multicenter cohort of people with HIV in the United States, March-December 2020. medRxiv 2021:2021.2010.2015.21265063.

23. Kitahata MM, Rodriguez B, Haubrich R, Boswell S, Mathews WC, Lederman MM, et al. Cohort profile: the centers for AIDS research network of integrated clinical systems. International journal of epidemiology 2008; 37(5):948–955.

24. Kozak MS, Mugavero MJ, Ye J, Aban I, Lawrence ST, Nevin CR, et al. Patient reported outcomes in routine care: advancing data capture for HIV cohort research. Clinical infectious diseases 2012; 54(1):141–147.

25. Crane HM, Kadane JB, Crane PK, Kitahata MM. Diabetes case identification methods applied to electronic medical record systems: their use in HIV-infected patients. Curr HIV Res 2006; 4(1):97–106.

26. Crothers K, Rodriguez CV, Nance RM, Akgun K, Shahrir S, Kim J, et al. Accuracy of electronic health record data for the diagnosis of chronic obstructive pulmonary disease in persons living with HIV and uninfected persons. Pharmacoepidemiol Drug Saf 2019; 28(2):140–147.

27. Levey AS, Stevens LA, Schmid CH, Zhang YL, Castro AF, 3rd, Feldman HI, et al. A new equation to estimate glomerular filtration rate. Ann Intern Med 2009; 150(9):604–612.

28. Eneanya ND, Yang W, Reese PP. Reconsidering the consequences of using race to estimate kidney function. JAMA 2019; 322(2):113–114.

29. Goff DC, Jr., Lloyd-Jones DM, Bennett G, Coady S, D’Agostino RB, Gibbons R, et al. 2013 ACC/AHA guideline on the assessment of cardiovascular risk: a report of the American College of Cardiology/American Heart Association Task Force on Practice Guidelines. Circulation 2014; 129(25 Suppl 2):S49–73.

30. Nunes D, Fleming C, Offner G, Craven D, Fix O, Heeren T, et al. Noninvasive markers of liver fibrosis are highly predictive of liver-related death in a cohort of HCV-infected individuals with and without HIV infection. Am J Gastroenterol 2010; 105(6):1346–1353.

31. Jain MK, Seremba E, Bhore R, Dao D, Joshi R, Attar N, et al. Change in fibrosis score as a predictor of mortality among HIV-infected patients with viral hepatitis. AIDS Patient Care STDS 2012; 26(2):73–80.

32. Kim H, Nance R, Van Rompaey S, Delaney J, Crane H, Cachay E, et al. Poorly controlled HIV infection: an independent risk factor for liver fibrosis. J Acquir Immune Defic Syndr 2016; 72(4):437–443.

33. Zou G. A modified poisson regression approach to prospective studies with binary data. American journal of epidemiology 2004; 159(7):702–706.

34. Hansen BB. The prognostic analogue of the propensity score. Biometrika 2008; 95(2):481–488.

35. Arbogast PG, Ray WA. Performance of disease risk scores, propensity scores, and traditional multivariable outcome regression in the presence of multiple confounders. American journal of epidemiology 2011; 174(5):613–620.

36. Tadrous M, Gagne JJ, Stürmer T, Cadarette SM. Disease risk score as a confounder summary method: systematic review and recommendations. Pharmacoepidemiology and drug safety 2013; 22(2):122–129.

37. Usman MS, Siddiqi TJ, Khan MS, Patel UK, Shahid I, Ahmed J, et al. Is there a smoker’s paradox in COVID-19? BMJ Evidence-Based Medicine 2021; 26(6):279–284.

38. Popkin BM, D. S, Green WD, Beck MA, Algaith T, Herbst CH, et al. Individuals with obesity and COVID-19: A global perspective on the epidemiology and biological relationships. Obesity Reviews 2020; 21(11):e13128.

39. Gregg EW, Sophiea MK, Weldegiorgis M. Diabetes and COVID-19: Population Impact 18 Months Into the Pandemic. Diabetes Care 2021; 44(9):1916–1923.

40. Boulle A, Davies M-A, Hussey H, Ismail M, Morden E, Vundle Z, et al. Risk factors for COVID-19 death in a population cohort study from the Western Cape Province, South Africa. Clinical infectious diseases: an official publication of the Infectious Diseases Society of America 2020.

41. Zheng H-Y, Zhang M, Yang C-X, Zhang N, Wang X-C, Yang X-P, et al. Elevated exhaustion levels and reduced functional diversity of T cells in peripheral blood may predict severe progression in COVID-19 patients. Cellular & Molecular Immunology 2020; 17(5):541–543.

42. Aljawharah Alrubayyi EG-M, Emma Touzer, Dan HameiriBowen, Jakub Kopycinski, Bethany Charlton, Natasha Fisher-Pearson, Pierre Pellegrino, Laura Waters, Burns Fiona, Sabine Kinloch-de Loes, Lucy Dorrell, Sarah Rowland-Jones, Laura McCoy, Dimitra Peppa. Characterization of SARS-CoV-2 Specific Responses in People Living with HIV. In: Conference on Retroviruses and Opportunistic Infections. Chicago, IL; 2021.

43. Halkitis PN. Managing the COVID-19 Pandemic: Biopsychosocial Lessons Gleaned From the AIDS Epidemic. Journal of Public Health Management and Practice 2021; 27.

44. Quinn KG, Walsh JL, John SA, Nyitray AG. “I Feel Almost as Though I’ve Lived This Before”: Insights from Sexual and Gender Minority Men on Coping with COVID-19. AIDS and Behavior 2021; 25(1):1–8.

45. Griffith GJ, Morris TT, Tudball MJ, Herbert A, Mancano G, Pike L, et al. Collider bias undermines our understanding of COVID-19 disease risk and severity. Nature communications 2020; 11(1):1–12.

46. Brown LB, Spinelli MA, Gandhi M. The interplay between HIV and COVID-19: summary of the data and responses to date. Current Opinion in HIV and AIDS 2021; 16(1):63–73.

47. Serpas DG, García JJ. Allostatic Load and the Wear and Tear of the Body for LGBTQ PoC. In: Heart, Brain and Mental Health Disparities for LGBTQ People of Color: Springer; 2021. pp. 41–52.

48. Amirkhan JH. Stress overload in the spread of coronavirus. Anxiety, Stress, & Coping 2021; 34(2):121–129.

49. Wakeel F, Njoku A. Application of the Weathering Framework: Intersection of Racism, Stigma, and COVID-19 as a Stressful Life Event among African Americans. In: Healthcare: Multidisciplinary Digital Publishing Institute; 2021. pp. 145.

50. Gil RM, Freeman T, Mathew T, Kullar R, Ovalle A, Nguyen D, et al. The LGBTQ+ communities and the COVID-19 pandemic: a call to break the cycle of structural barriers. The Journal of infectious diseases 2021.

